# Patient-public involvement (PPI)-led qualitative analysis of >400 Inflammatory Bowel Disease patient responses in United Kingdom: An independent patient-led review to provide insights to improve care and research

**DOI:** 10.1101/2025.03.19.25324163

**Authors:** Molly J Halligan, Aerin E Thompson, Destiny Docherty, Patricia Kelly, Emma Pryde, Cher Shiong Chuah, Rebecca Hall, Gwo-tzer Ho

## Abstract

**Background:** This paper investigates a patient-led approach to research on wellbeing in individuals with Inflammatory Bowel Disease (IBD). Traditionally, Public and Patient Involvement (PPI) groups have contributed to the design of clinical research but less often to the analysis and reporting of findings. This study is wholly led by a patient group with no direct input from clinicians, thus presenting an entirely novel and unique patient-centric view.

**Methods:** This report draws on data from a Wellbeing Survey led by the Edinburgh IBD Science team as part of the MUSIC IBD cohort study (www.musiscstudy.uk) with over 1,375 IBD respondents over two time periods in 2023 from United Kingdom. The PPI group utilised high-level topic analysis and their own lived experience of IBD to explore the 415 free-text survey responses. Regular discussions allowed the team to reflect on patient narratives and generate findings collaboratively. PPI members contributed to both the structure and content of the final write-up, utilising their diverse backgrounds and skillsets.

**Results:** The analysis provided in-depth exploration of several key themes affecting wellbeing in IBD patients. Of interest, the PPI group discussed and explored themes such as ‘what does remission mean?’, access to care, expectations of self-management, mental and women’s health. The patient narratives highlighted the variability of IBD experiences, the interconnectedness of these issues, and the importance of holistic, patient-centric approaches to care. The findings emphasise the necessity for improved support, both within and beyond healthcare settings.

**Conclusion:** This patient-led research approach demonstrates that allowing patients to lead in analysis (‘taking the reins’) and reporting provides deeper and impactful insights into IBD experiences. By integrating patient perspectives, this study advocates for a patient-dominant approach to research and care, which can improve outcomes and support ways to address the complexities of living with IBD. The model highlights challenges and benefits of this approach, serving as a foundational template for future patient-led collaborations, in addition to the immediate impact of patients’ wellbeing from shared experiences, to educating clinicians and people without IBD about the impact of IBD on patients’ lives.

## Clinician’s summary: The origins of our patient-led project

Patient-public involvement (PPI) is now essential to all fields of research including fundamental science. From all aspects, PPI improves the quality, safety and impact of our research^1^. Many patients also derive direct benefits of taking part in PPI and gain a sense of agency in the research and their conditions as well^2^.

Collectively in the field of research, we are learning quickly in how to improve the quality of PPI. Historically, the researchers play a leading role. By asking patients to provide their input which are then processed and directed towards a particular goal. More recently, a ‘co-production’ approach, where patients take an equal equity to develop a PPI effort with researchers is increasingly used^3, 4^.

In our research that spans fundamental science and wellbeing in IBD, 1, 375 IBD patients have recently taken part in 2023; with more than 400 detailed responses provided in all areas relating to the impact on IBD on their lives. **Here, we are presented with a unique opportunity to allow our PPI group to lead (‘take the reins’).** This was conceived from discussions within our PPI group and places all the onus on our PPI group to review, analyse and present this IBD patient-reported dataset. In short, a wholly patient-led review with the responsibility to appraise all our IBD respondents’ views. Conceptually, this work is primarily produced **‘by the patients, on data provided from the patients, and for the patients.’**

The result of this collective PPI effort is radiantly impactful and presents insights that are entirely novel in their perspectives and nature. As a clinician and a researcher, I have learned a lot from this PPI-led work and hope that this work will be helpful for future PPI approaches, patients and all the stakeholders involved in improving the care of our IBD patients.

## Introduction

This paper explores the role of the patient in delivering research related to wellbeing living with Inflammatory Bowel Disease (IBD). This paper will explore a patient-dominant model and approach to research: where patients lead in defining the topic areas, setting the agenda, and communicating information to the target populations: the IBD community, clinicians and researchers. The following paper has been written from the perspective of people with lived experience of IBD.

Within IBD clinical care, there has been a shift to understand experience beyond inflammatory markers (bowel frequency, calprotectin tests, and blood samples). IBD clinicians now want to know about fatigue levels, daily stresses, and how a patient personally feels their IBD is being managed. These patient-centric interactions within healthcare are increasingly being transferred to clinical research, which seeks to take a more holistic approach to understanding issues for people with IBD often in collaboration with Public Patient Involvement (PPI) groups.

PPI groups are where patients, members of the public and researchers or clinicians co-produce solutions and outputs to improve services. These groups allow patients to be more involved in decision making and developments related to their care. PPI groups are routinely asked for their invaluable insights in the design or development stages of clinical research, such as reviewing participation sheets or final research papers, but in analysis and writing up results they often assume a less dominant role. This is primarily due to the technical knowledge that is often required to analyse scientific or specialised information. However, as research continually focuses on and values the patient’s perspective, outputs are no longer restricted to understanding clinical results, such as immune-cell function or biomarkers. Instead, responses from patients, provided in their own words, can be analysed to aid a deeper understanding of these clinical results. PPI group members with lived experience of a condition can provide invaluable knowledge to aid this analysis and write-up of results.

The aims of this paper are to explore the value of the patient voice in understanding and analysing experiences of IBD in relation to Wellbeing by:

1. Understanding the priorities around research and wellbeing from the perspective of survey respondents and the PPI group.
2. Understanding how patients analyse, present and communicate patient reported concerns of wellbeing and experience with IBD.
3. Understanding how to make this process more transferrable for future projects (specifically, how to make this appealing to clinical researchers).

## Methodology

The process for this paper began with the MUSIC IBD study (www.musicstudy.uk; ethical approvals by East Scotland Ethics Committee REC 19/ES/0087 and R&D No: 2019/0325), a clinical study that aimed to look at Mitochondrial DAMPs as mechanistic biomarkers of gut mucosal inflammation in adults and children. The MUSIC IBD clinical team ran a Wellbeing Survey to compare patient-reported outcomes (PROs) of people with IBD to those without this condition (accessible via https://forms.ocice.com/e/aJWUEC4UUN). The survey captured information from over 1,375 IBD patients from two time points (January – March and June – September 2023) in an anonymous manner. This study included 40 closed questions, where respondents were asked about bowel habits, fatigue, mental health and general wellbeing. At the end of the Wellbeing survey, respondents were asked whether they had ‘any further comments that you feel are important and have not been addressed?’. This resulted in 415 free-text comments from respondents with IBD. A parallel research paper that is focused on IBD fatigue is available on https://doi.org/10.1101/2025.01.18.25320777 on medRxiv.

Most respondents to the survey answered this question by sharing insights into their experiences of living with IBD to express what was important to them, often providing more detailed context to the closed questions in the survey. As the PPI group, clinicians and researchers discussed these free-text responses, it became evident that insights from the PPI members with lived experience of IBD were strengthening the interpretation of these comments. These PPI members were then given the opportunity to lead on the analysis and write-up of the free text responses.

As well as their experience of IBD, the PPI representatives brought a range of skills from their academic and creative backgrounds, including social research and qualitative analysis, immunology and inflammation-based scientific knowledge and experience of prior PPI work. This allowed for this type of analysis and style of report to be developed without external guidance.

The following approach was agreed and carried out by the PPI group:

a) **Analysis:** High-level topic analysis was conducted to draw out main priority topic areas for the survey respondents with IBD using colour coding in Excel. These initial topic areas were then reviewed by all members of the PPI group and used to guide the structure of the paper.
b) **Topic Areas:** The PPI group agreed that representation of all topics, even those where only a few survey respondents mentioned it’s importance (e.g. age), should be considered to capture the variation of experience with IBD. It was agreed that PPI members would lead on 3-4 individual topic areas to encourage collective leadership and result in a well-rounded paper informed by multiple perspectives.
c) **Discussion:** It was agreed that the PPI group would discuss each topic area together over regular Teams calls (20 minutes per topic). Seven meetings were held to discuss themes. PPI members would explore the free text comments relevant to their topic, select a few that resonated with them or made them reflect differently on their experience, and bring their initial thoughts to the meeting. The discussions would focus on why this topic may have been important for survey respondents and the PPI groups’ similarities and differences in experience to expand on the topic area further. The topic lead would take notes throughout the meeting, which would be used to guide their write up for the paper.
d) **Writing:** It was agreed that PPI members would write up their own topic for the report to allow the paper’s narrative to develop naturally. To utilise the wide skill set of the PPI group, PPI members were encouraged to use any additional sources they felt were relevant to develop their topic (e.g., including literature or sources from journals or charities^1^). As the paper developed it became clear that the overall style of writing was academic, reflecting the PPI members who came from an academic background. This presented challenges for PPI members who were less familiar with this style and engaging with scientific journals or jargon. After producing an initial draft, the approach was adjusted and individuals with less familiarity with academic writing took on a more review-oriented role, providing feedback and further input to their sections as they were refined. As the process slowed and fewer contributions were made, the narrative was streamlined and other sections outside of the results (e.g., methodology) were written. The final version was peer reviewed by the PPI group before being shared and reviewed by the clinical research team.

By having multiple contributors involved in the process, the PPI group developed a paper that explores a deeper understanding of IBD and wellbeing that might not have been possible if interpretation and analysis was conducted by clinical researchers without IBD. However, the 400+ free text comments and the experiences shared by the PPI group highlight that no two experiences of IBD are the same and therefore, the experiences discussed in this paper may not be representative of all people with IBD.

Two potential risks of developing a patient-led paper are that some of the participants may become unwell during the process due to the unpredictable nature of IBD or be unable to attend all discussions due to busy schedules. To mitigate this several actions were taken: 1) additional notes were taken during every discussion as a back-up copy in case of absences during the writing stage; 2) dividing topics among five PPI members to distribute workload; and 3) provide opportunities for PPI members to input their experiences and opinions ‘offline’ via written submissions for missed meetings. Finally, the group shared resources with each other to support wellbeing as often some topics were sensitive to discuss.

Quotation marks have been used when referring to responses from the survey. For the purposes of anonymity and this condensed version of the paper, PPI member’s individual experiences and survey respondent comments have been merged. We highlight issues from survey responders and PPI group members in respective text boxes.

## Findings

The following subsections are based on the high-level topic areas produced from the survey responses and further explored by the PPI group. A decision was made early in the process to omit the number of respondents that mentioned each topic area. It was agreed that quantifying the experiences distracted from the importance of the topic area to consider for further research. In addition, the complexity and variation in experience of IBD suggests that respondents may have experience of more than one topic, even if they did not mention it in their response to the question.

To give some indication of importance and prioritisation of the impacts on Wellbeing with IBD, the topics discussed below are ordered by the frequency they were mentioned in the survey results, e. g. the impact of IBD on quality of life was mentioned the most throughout survey responses and ageing with IBD was mentioned the least.

## Quality of Life

*“I think it would be important for people to be more aware of how many areas of your life IBD aJects” –* survey respondent.

Quality of life (QoL) in IBD patients encompasses physical, emotional, and social wellbeing which is ultimately determined by how much the disease impacts an individual’s life, what is important to them, and how well the disease is managed. The QoL areas detailed here will be further examined throughout this paper however, it is important to explore this theme directly, as respondents spoke about the significant ways in which IBD impacts QoL overall.

### The survey responses and PPI group noted

- The mental and physical impact of IBD symptom management on daily activities, such as planning ahead to know where the toilets are when outside the home, eating bland food to avoid aggravating the gut, and staying on top of medication.
- The psychological impact of IBD due to the chronicity of the disease, threat of flare-ups and associated poor mental health.
- Challenges around being able to socialise due to the unpredictability of IBD compounded by fears of being misunderstood or judged for not engaging with events or social activities.
- Lack of support or adjustments from employers and in education settings which makes it difficult to cope with the relapsing and intermittent nature of the disease.

Throughout the survey responses, an overarching theme of “surviving, not living” came through. Overall enjoyment in life and the ability to carry out day-day activities are inhibited by the symptoms and limitations imposed by IBD. While this is specific to an individual, the burden that comes with living with a chronic disease is common to patients. However, as will be explored through each of the following sub-headings, when the right management methods, support, and understanding from others are found then the burden of IBD on QoL and wellbeing can be reduced dramatically.

## Access to care

*“Having an eJective and approachable IBD team has been a lifeline for me…” –* survey respondent.

Cost of and access to care was the second most prominent topic to emerge from the survey with respondents reporting on their experiences interacting with healthcare services and professionals.

- **The survey responses and PPI group noted:** Delayed access to the correct care.
- Not feeling as though medical professionals are listening properly to reported symptoms.
- Being given insufficient information or not fully understanding the information given to them about medication or IBD by healthcare providers. This was thought to be worse when transitioning from child to adult IBD services, where the care received in adult services is less intensive, and communication is not as often.
- When requiring care abroad or moving to new areas, people with IBD may require extensive knowledge of their own medical history to be able to receive care from new healthcare providers who may not have access to their original medical records. This is especially challenging if someone has multiple conditions or must speak to non-specialised healthcare providers about their IBD medication or symptoms.
- The time and financial cost of navigating access to IBD-related healthcare, where managing multiple medications and interactions with healthcare providers can be challenging.

Overall, the PPI group and respondents felt that these issues resulted in people with IBD feeling as though they need to manage their condition themselves. Improvements could be made through better communication between healthcare professionals and people with IBD, as well as realistic solutions to accessing appropriate care such as more local support or ‘care in the community’ approaches.

It is important to note that respondents to this survey and PPI members also wished to comment on times where their access to care and treatment has worked for them and expressed gratitude for the services they received. In addition, the PPI group agreed that public and patient engagement gives a unique insight into IBD beyond the patient perspective and allows them to have a greater appreciation of the pressures that the NHS are under, as well as gratitude for the work and research that goes on behind the scenes.

## What does ‘remission’ mean?

*“Despite being in remission, I have daily struggles and pain due to these issues so remission isnt really an accurate description”* – survey respondent.

The concept of being ‘in remission’ varied widely among respondents, with many highlighting the ambiguity and confusion surrounding this term. The most common experience of remission reported by survey respondents and the PPI group challenged the concept of remission as recovery.

- **The survey responses and PPI group noted:** The confusion about the term ‘remission’, where it can be applied to someone who has no active inflammation or symptoms, someone who has some mild and less severe symptoms, and someone who has no active inflammation but still presents with continuation of typical IBD symptoms.
- The frustration and confusion, as patients find themselves labelled as “well” despite continuing to live with symptoms and feeling as though these symptoms are not properly acknowledged by their healthcare professionals.
- The broader implications of labelling a person as ‘in remission’ may mean that people with IBD feel as though they are somehow responsible for their symptoms continuing, or they are being perceived as over-exaggerating symptoms by clinicians, family and friends.
- That the diagnosis of ‘remission’ may mean there are no other options available to solve their persisting symptoms which are impacting their wellbeing and QoL.

Overall, the PPI group believes that there are communication and language issues around the term ‘Remission’ and that it would be useful to develop new language to describe these different states of ‘wellness’. This would not only have an impact on assessing and measuring patient clinical and self-reported outcomes but also improve the broader implications that the term ‘remission’ can have on a person’s life, as described above.

## Mental Health

*“I feel very anxious about going out & doing normal activities. I know stress is a major factor to trigger my symptoms of IBD and trying to control my stress levels is sometimes hard” –* survey respondent.

Many respondents mentioned mental health, particularly stress, anxiety, and depression, in relation to their wellbeing with IBD. These were often related to trying to manage the condition outside the home, aftereffects of surgery, and anxiety around the potential or current condition of flaring. However, the PPI group and respondents also shared experiences where their clinician’s choice in treatment or change in approach after listening to their concerns removed anxieties and improved general wellbeing.

- **The survey responses and PPI group noted:** The unpredictability of living with IBD and how this challenges a person’s ability to carry out daily activities or be able to work can have an adverse impact on their mental health.
- Environments, such as a stressful job or university exams, can cause poor mental health outside of having IBD and can often not be controlled. However, the PPI group also felt it was important to note that poor mental health itself may not cause a flare, and the onus should not be on the patient to ‘just relax’ or ‘manage stress better’ as a solution or preventative measure to reduce IBD flares as this is not always possible.
- The side effects of medication to help manage IBD symptoms, such as steroids, can result in mood changes and depression. This may influence some people with IBD to avoid these treatment options to reduce the impact on their mental health.
- The anxiety and uncertainty around starting new or invasive treatments, complications of treatments or surgery, and whether they will be able to tolerate the pain or side effects. Also, the mental and physical perseverance that is needed when treatment options do not work.
- Changes to their body image and the impact of stoma surgery, both topics discussed in sub-headings below.
- The impact of social isolation where understanding and support from others is needed over long periods of time but may be exhausting to provide. Equally, not being able to commit to events or cancelling social interactions last minute due to the unpredictability of IBD can contribute to feelings of loneliness.
- The cost of accessing private mental health treatment, where free alternatives are not offered as part of IBD care often due to waiting lists and staff shortages. This will not be an affordable option for all patients, so many will have to wait for free treatment and extend their time with unsupported poor mental health.

Overall, the impact of poor mental health and IBD is complex and requires further exploration in research, particularly in cases where the patient cannot control the factors or environments causing additional stress and potentially exacerbating IBD symptoms. Based on the survey responses and PPI discussion there is a need for appropriate and accessible mental health support to be commonly offered as part of IBD treatment.

## Extraintestinal Manifestations of IBD symptoms

*“I feel the discomfort, stress and physical internal body pressure caused by bloating and/or by constipation is completely underestimated as a problem by clinicians”* – survey respondent.

Respondents mentioned ‘non-typical’ IBD symptoms, referring to symptoms other than loose stools and blood in the stools, that they felt were important to highlight as impacting their experiences and wellbeing with IBD. Examples of extraintestinal manifestations of IBD symptoms included constipation, vomiting, pain, bloating and other symptoms respondents believed are explicitly linked to their IBD.

- **The survey responses and PPI group noted:** The non-typical IBD symptom which was mentioned the most was constipation, which people felt was often overlooked despite being problematic.
- The second most mentioned non-typical IBD symptom was pain generally and, more specifically, abdominal pain. The PPI group felt that pain is not comprehensively monitored despite being experienced by most patients and it can be easily dismissed as being related to other things, such as poor diet, menstruation pain or appendicitis. This can be particularly problematic pre-diagnosis and also cause confusion for people with IBD when they are trying to identify and report to clinicians what is and is not IBD.
- Treatments that are usually aimed at tackling “physical gut symptoms” without consideration for other issues which are equally important to treat. Other issues mentioned include, joint pain, mouth ulcers, erythema nodosum (a skin condition which causes red nodules usually on the shins), issues with eyes such as dryness, and ectopic Crohn’s disease (Crohn’s located in the vulva).

Respondents highlighting these symptoms may indicate that it was not considered a typical IBD symptom when the survey was created. As extraintestinal manifestations of IBD are common among people with IBD, researchers should consider exploring these further in future IBD studies. The PPI group discussion and survey respondent comments also highlight the clear need for stronger communication between clinicians and patients in order for people with IBD to receive the correct care and medication. This may require support from a multidisciplinary team depending on where these additional IBD symptoms are located.

## Impact of Additional Conditions

*“when there are comorbidities things get very challenging.. medications get even more confusing and capacity to focus on healing IBD gets very hard as it’s hard to manage everything all at once..”* – survey respondent.

Many respondents spoke about additional conditions that they did not explicitly link to their IBD but still felt that it impacted their life with IBD, such as COVID, fibromyalgia or other autoimmune disorders.

- **The survey responses and PPI group noted:** The importance of asking about other short or long-term conditions when conducting IBD research as responses may be influenced by other issues.
- The level of knowledge required to ascertain (often guessing) what is and what is not an IBD related issue and reporting this back to clinicians to acquire the correct support.
- The challenges of coordinating care where a person may have a range of conditions to manage at the same time. This may include engaging with a variety of different healthcare professionals across multiple disciplines and relying on communication between these teams to consider potential drug interactions, additional side effects, and adherence challenges. This level of coordination can be an added layer of stress to a patient’s treatment journey.

Overall, this topic highlights the importance of acknowledging other short or long-term conditions that introduce unique challenges to the management of IBD. Comprehensive coordination of care, particularly for people with dual pathologies or multiple conditions, is essential for addressing challenges and to provide effective IBD management. Ensuring that all aspects of health are considered, alongside communication and knowledge shared by the correct healthcare professionals for improved decision-making and patient wellbeing.

## Awareness and knowledge about IBD

*“I think it would be important for people to be more aware of how many areas of your life IBD aJects. It is not just a disease that makes you need the toilet a lot”* – survey respondent.

Lack of awareness and knowledge of the reality of IBD was mentioned by respondents of the survey as having a huge impact on their wellbeing, particularly influencing the support and compassion they receive from others.

- **The survey responses and PPI group noted:** The impact of stigma on a person’s experience of IBD and related wellbeing. IBD can be embarrassing, debilitating and misunderstood which can exacerbate existing stigmas and prevent people from disclosing their condition to others.
- Equally, the invisible nature of IBD means that people with IBD can appear visually well, but they may be experiencing a flare without noticeable symptoms. This can often make it challenging for others to understand a person’s needs.
- IBD is often not recognised as a “serious illness” by local authorities and workplace or education settings which reduces specific accommodations or support for people with an invisible illness. Examples include toilet accessibility, financial support and parking accessibility.
- Experiences of isolation due to lack of awareness about IBD symptoms and experience.
- The onus may be on the person with IBD to always educate others and that it is important for family/friends to do their research and increase their own knowledge about the condition themselves to lessen the burden on the person with IBD.

However, some respondents and PPI members spoke about the flexibility provided in hybrid or home working and that now employers seem somewhat more understanding about workplace adjustments since the COVID-19 pandemic. The PPI group, including clinicians and researchers without IBD, have also focused efforts on increasing awareness which include the release of the short film “Our lives with IBD” at the Edinburgh science festival. This short film explored IBD through the experiences of those living with it and the ongoing research by the clinical team. Opportunities to share knowledge about IBD also exist via charity networks, such as Crohn’s and Colitis UK, allowing people to share their experiences and strategies for coping with various issues.

Overall, the survey respondents and PPI group emphasised the need for more awareness of IBD, particularly around the invisible nature of IBD, which may help reduce isolation and increase overall support for a person with this condition. As shown above, this can be achieved by clinicians and researchers through creative and existing projects but needs to be accessible to and attended by those without IBD.

## Self-management of IBD

*“How to better manage oneself, keen to take responsibility for myself and understand what I can do to improve my condition”* – survey respondent.

Treatment and management of IBD involves a multifaceted approach including a range of medical interventions such as immunosuppressants, steroids and surgery. In addition to conventional medical treatments, self-management and additional methods of care have a significant role in IBD and can contribute to improving overall patient outcomes, as seen by survey respondents’ interest in learning about additional supportive methods.

### The survey responses and PPI group noted

- Lack of information about developing self-management options that can be tailored to specific patient needs, such as dietary adjustments or how to exercise safely with certain symptoms.
- The lack of mental health and psychological support. As discussed earlier, this would be influential in developing effective coping mechanisms and approaches to stress management.

The management of IBD through self-driven approaches and additional methods empowers patients to take a proactive role in controlling their health with support from healthcare professionals. This includes the correct support for optimising nutrition, safe and effective exercise through improved patient guidelines and plans, and self-led approaches to support mental health which may include stress management and relaxation techniques, therapy and counselling, or flexibility in an individual’s working environment. However, in the same way that IBD does not affect all patients identically, the resounding theme was every patient must approach additional methods of care individually.

## Women’s Health and Reproduction

*“IBD is contributing to heavy and painful menstrual cycle irregularities causing heavy periods which worsen IBD symptoms and vice versa”* – survey respondent

Women’s Health and Reproduction was an emerging topic that some respondents felt was important to them and should be examined further.

- **The survey responses and PPI group noted:** Lack of information about developing self-management options that can be tailored. The negative influence of menstruation and menopause on IBD and/or bowels generally. This includes changes in menstruation patterns, abdominal pain, fatigue, headaches, and increased bowel changes. Equally, issues with low iron which makes symptoms like fainting and dizziness worse during menstruation.
- That IBD and menstruation related bowel urgency and pain can often feel similar when located in the lower abdomen, which causes confusion about knowing when issues may be IBD related or not.
- Concerns about fertility, maintaining remission during pregnancy and the impact of pregnancy on IBD symptoms or flares. This may indicate that this topic is not widely discussed or there is not enough information readily available to people with IBD.
- Feeling limited in relation to contraception due to worries about extra hormones or side effects in addition to IBD medication, such as steroids, which may make IBD symptoms or mental health and wellbeing worse.
- Worries about having a flare during pregnancy and whether maternity doctors and nurses have enough knowledge about IBD to support properly.
- Managing IBD flares, miscarriages, and resulting mental health challenges simultaneously can be deeply traumatic, and this struggle often intensifies the emotional toll of living with IBD, as it not only affects their physical health but also their sense of identity and future aspirations, including family planning.

Although the Crohn’s and Colitis website offers information about fertility, pregnancy, and breastfeeding, the PPI group felt that the impact of IBD on pregnancy and the side effects of IBD medication on pregnancy isn’t well known or widely discussed outside these resources. Timely discussions about women’s health and reproduction, including proactive conversations and decision-making about treatment options between clinicians and women who want to conceive, and communication with maternity teams about IBD to inform them of potential scenarios and specific support is required.

## Exhaustion and Fatigue

*“Fatigue is underrated. I cannot remember the last time I had energy”* – survey respondent.

It is important to note that some core survey questions were aimed directly at understanding fatigue. Despite the fact it was mentioned in the survey, these respondents re-emphasised the notion that “fatigue is underrated” in its effects on daily life but that it can “often [be] one of the biggest side effects of IBD”.

- **The survey responses and PPI group noted:** Treatment options for fatigue are not always effective
- Mental and physical fatigue are often not considered severe symptoms of IBD and there is a risk of looking lazy by employers or school tutors.
- The unpredictable nature of fatigue and how difficult it can be to plan social activities, plan for work or academic responsibilities, and having to disclose IBD to fully express the effects of fatigue.

The PPI group discussed the benefits of organising social life along with work at a slower pace, for example planning social engagements, sports or activity, and working hours to be spread out more evenly across the week/month. The group also spoke about being open with peers and colleagues, so in the circumstance of having to cancel events, or work from home due to fatigue, people may be more understanding of their condition. However, these approaches are not always possible, and that more understanding and compassion from people without IBD is required.

When looking at methods of care for fatigue, many online sources advice is often limited or not written from a patient aspect: ‘eat better or take supplements’, ‘get tested for anaemia’, or simply ‘get a better sleep schedule’. It is important to note that some treatments for fatigue, such as B12 injections or iron infusions, do work for certain patients and such treatment options should be an ongoing discussion between the clinician and the patient. However, survey respondents and the PPI group have stated how tackling fatigue is not as straightforward as improving sleep schedules or taking more vitamins and minerals. It is clear that further research, such as surveys focusing on patient reported outcomes, is necessary to support individual experiences of fatigue.

## Body Image

*“I struggle with constipation and the pain and discomfort leave me feeling unattractive generally” –*survey respondent.

More recently, improving negative body image is increasingly part of mental health awareness in society. Despite IBD being considered an invisible illness, the impact of the condition, particularly stomas, on a person’s body image can leave individuals feeling uncomfortable in their own skin.

- **The survey responses and PPI group noted:** Poorer sense of body identity and body image with IBD which can influence social engagement, stylistic choices and romantic aspects of life.
- Issues such as mouth ulcers or steroid treatments causing facial swelling and rashes on the skin, as a side effect of IBD or medication, making people feel self-conscious. To further this, IBD can cause bloating and rapid weight loss, which again removes the sense of “autonomy” or “possession” over one’s own body and identity, especially when making adjustments using diet and exercise, which may be limited due to IBD.
- People may opt for baggier, more patterned designs in clothing to accommodate and hide stomach bloating or stomas (discussed in the next sub-heading) but these may be clothing that does not align with their fashion identity.
- Due to IBD symptoms that have a physical effect on the body, people with IBD may avoid romantic engagement with others, such as dating or sex, which contributes to feelings of loneliness and poorer perception of self.
- Some might exhibit avoidance behaviours by refusing medications which would usually change body image, such as gaining weight from steroids and having what is known as a moon face.

Further research and guidance on changes to body image and perception of self when diagnosed or living with IBD would be beneficial. Clinicians should consider discussing this topic with patients more frequently in consultations, especially if medication adherence is impacted, and may wish to refer the patient onto further support from mental health professionals.

## Experience with Stomas

*“My stoma has gave me life back however it does come with it’s [own] challenges for example fear of leaks. However, I am in a lot less pain and able to do things I could not when I was in a flare up like social things. My quality of life is a lot better with my stoma”* – survey respondent.

When IBD is not well controlled by medication, sometimes the only option is to have surgical intervention and create a stoma. A stoma is where the end of the bowel is diverted out through the abdominal wall and sits just outside the skin of the abdomen This requires wearing an ‘appliance’, ‘bag’ or ‘pouch’ to collect the waste from the stoma. Nobody who considers stoma surgery does so lightly. It is often the only option when in a life-threatening situation due to complications from flaring IBD.

- **The survey responses and PPI group noted:** Being scared about the prospect of having to have stoma surgery.
- The development of different management strategies to cope with the challenges of living with a stoma, such as leaks or rashes.
- Misconceptions and fears of having a stoma, for example, in some cases, a stoma does not necessarily mean that a patient no longer has IBD or related symptoms to manage which can be confusing for patients.
- Respondents described how going through stoma surgery can change their self-perceived body image. This may include fears around how someone’s partner will view them or challenges of dating with a stoma.
- As mentioned in previous chapters, IBD is an invisible condition, but surgery is a visible reminder of the condition that people may feel they have to hide.
- Some patients are given little to no mental health support in adjusting to having a stoma and are left to try and figure things out on their own.

However, having a stoma can also really improve a patient’s quality of life. Respondents to the survey spoke about how stomas have improved their quality of life and that they are able to take part in social activities without being burdened by their usual IBD symptoms. When discussing potential stoma surgery with patients it’s important to acknowledge this trade-off.

As mentioned in a previous subheading, improving mental health support and awareness of IBD is considered important. However, mental health support and awareness of IBD specific to stomas is required as the respondents and PPI group agreed that it is a unique experience which is difficult to understand without going through it yourself.

## Age

*“My biggest fear is when I get older and not being able to control my bowel”* – survey respondent.

Age in relation to IBD refers to how IBD symptoms and severity can change over time as well as the worries and concerns patients have about aging and having IBD.

- **The survey responses and PPI group noted:** The misconception that someone young is considered ‘healthy’ due to the invisible nature of the condition. This view can make it challenging for individuals to seek help and to talk about their condition as well as to come to terms with being young with a chronic illness.
- Fears around getting older with IBD and what this might mean for bowel control or the impact on additional conditions. It is often difficult to know how IBD will progress over a person’s lifetime which makes treating IBD challenging and makes predicting how someone’s experience with IBD may affect them in the future difficult.

IBD and age is a topic which is considered and heavily studied in terms of the prevalence of IBD among different age groups but less so about the impact of age on experiences with IBD. Strategies have been used to help individuals under paediatric care to feel less isolated, to understand their condition more and to direct research in a way that is driven by patients, such as the “Let’s talk about research” paediatric IBD event held at the Royal Hospital for Children and Young People (<u>CIR hosts patient and family day exploring paediatric IBD | Centre for Inflammation</u> <u>Research</u>). This event was hosted by the centre for inflammation research in Edinburgh and allowed young people and their families to learn more about IBD as well as brought together scientists, research clinicians and patients. Further research into age and experiences of IBD is needed, especially as it influences nearly all topics discussed in this paper and wellbeing overall, and people with IBD may appreciate opportunities to learn more about variation of experiences with IBD based on age.

## Discussion

This paper has explored the priorities around research and wellbeing that are important to continue to improve experiences and outcomes for people with IBD. Equally, this paper acts as a reminder that any efforts to improve the lives of people with IBD, whether that is examining particular cells or organising patient-events, have the ability to positively impact one, if not more, of the topics discussed. This paper also acts as an example that for research to be effective, it cannot be done without the invaluable knowledge of the patients themselves.

It is important to note the interconnectedness of the issues raised in this paper and that there are commonalities between the topic areas, even when individual variation in experience occurred. These included concerns about support from wider circles outside of healthcare settings (family, friends, university or work), reviewing language around IBD, fears about social isolation and loneliness, the responsibility to manage one’s own IBD, improved communication from healthcare professionals, and seeking expansion of support from mental health and multidisciplinary teams. It is important to consider these interconnected themes to understand the complexity of managing IBD as well as develop better holistic and patient-centred care plans.

Equally, patient-reported outcomes are not only important for clinicians to understand but also expands patients’ knowledge of their or others’ experiences of IBD when made accessible to a non-technical audience. Developing this paper allowed the PPI group to learn more and gain new perspectives on IBD and the variation in experiences.

The PPI group found analysing, presenting and communicating patient reported outcomes an ambitious and rewarding experience. The range of skills, including academic backgrounds, created the template for this paper, however other PPI groups with members who have different skills may produce different outputs. The process for this paper was created from the ‘ground-up’. Benefits of the process were discussed, such as the opportunity to lead topic areas, the number and length of meetings to discuss topic areas (although frequency may need to be adapted depending on the groups preferences), the opportunity to learn from other experiences, being able to delve into the complexities of each topic area, and give a voice to the respondents from the survey. Reflections on how to make the process smoother included delegating tasks based on skill sets, creation of a template with resources to guide clear communication, and more focus on the impact of the work earlier on in the process. The group also reflected on being entirely patient led for the discussion and writing process. All members enjoyed that discussions were only attended by the PPI members who had IBD. Some members felt that additional support may have been beneficial during the writing process, however all liked the freedom to initially write without clinical input.

## Conclusion

This experience of patient involvement as a collaborative process has been rewarding for the PPI group, allowing them to provide a voice to the survey comments and produce something meaningful. Some respondents to the survey answered the free-text question by saying how pleased they are to fill out a survey focusing on patient-reported outcomes, highlighting how meaningful the survey was to the people who filled it out. However, despite the group’s best efforts to capture the variation, there will always be more to understand.

Although it is rewarding to have the patient involved in the decision-making process around research and to have their voices represented in this paper, the group would like to see this work steer conversations about conducting IBD research in new ways. In particular, around the clinical-research community where co-produced and patient-led models are perhaps not frequently seen as beneficial or worthwhile.

The PPI group would like to thank the survey respondents for giving up their time and detailing their experience of wellbeing with IBD for the purposes of this study. The group would also like to thank Professor Gwo-Tzer Ho for allowing them to ‘take the reins’ of this research and produce work that platforms the voice of people with IBD.

## Appendix

All patient authors provided consent to sharing their identities and background of their IBD conditions.

## Contributions

MJH, AET, DD, PK and EP are IBD patients and performed the review of available data, manuscript writing and presentation of findings. MJH coordinated the development of this project. RH, CSC and GTH are IBD clinicians and provided access to the patient datasets. GTH provided the Sponsorship and overview of this project. GTH and MJH conceived the idea of this patient-led project. This manuscript is wholly produced by the patient group with no direct input from the IBD clinicians except for the abstract, clinician summary and formatting of the paper.

## Data Availability

All data produced in the present study are available upon reasonable request to the authors.

## Acknowledgements

We thank all the IBD patients who took part in our study. Crohn’s Colitis UK, Catherine McEwan Foundation and Cure Crohn’s Colitis for supporting our work.

## Footnotes

1 These have been removed for this condensed version of the report. Please see the full version for expanded detail.

